# Reducing the equity gap in under-5 mortality through an innovative community health program in Ethiopia: An implementation research study

**DOI:** 10.1101/2023.05.30.23290706

**Authors:** Laura Drown, Alemayehu Amberbir, Alula M. Teklu, Meseret Zelalem, Abreham Tariku, Yared Tadesse, Solomon Gebeyehu, Yirdachew Semu, Jovial Thomas Ntawukuriryayo, Amelia VanderZanden, Agnes Binagwaho, Lisa R. Hirschhorn

## Abstract

**Background:** The Ethiopian government implemented a national community health program, the Health Extension Program (HEP), to provide community-based health services to address persisting access-related barriers to care using health extension workers (HEWs). We used implementation research to understand how Ethiopia leveraged the HEP to widely implement evidence-based interventions (EBIs) known to reduce under-5 mortality (U5M) and address health inequities.

**Methods:** This study was part of a six-country case study series using implementation research to understand how countries implemented EBIs between 2000-2015. Our mixed-methods research was informed by a hybrid implementation science framework using desk review of published and gray literature, analysis of existing data sources, and 11 key informant interviews. We used implementation of pneumococcal conjugate vaccine (PCV-10) and integrated community case management (iCCM) to illustrate Ethiopia’s ability to rapidly integrate interventions into existing systems at a national level through leveraging the HEP and other implementation strategies and contextual factors which influenced implementation outcomes.

**Results:** Ethiopia implemented numerous EBIs known to address leading causes of U5M, leveraging the HEP as a platform for delivery to successfully introduce and scale new EBIs nationally. By 2014/15, estimated coverage of three doses of PCV-10 was at 76%, with high acceptability (nearly 100%) of vaccines in the community. Between 2000 and 2015, we found evidence of improved care-seeking; coverage of oral rehydration solution for treatment of diarrhea, a service included in iCCM, doubled over this period.

HEWs made health services more accessible to rural and pastoralist communities, which account for over 80% of the population, with previously low access, a contextual factor that had been a barrier to high coverage of interventions.

**Conclusions:** Leveraging the HEP as a platform for service delivery allowed Ethiopia to successfully introduce and scale existing and new EBIs nationally, improving feasibility and reach of introduction and scale-up of interventions. Additional efforts are required to reduce the equity gap in coverage of EBIs including PCV-10 and iCCM among pastoralist and rural communities. As other countries continue to work towards reducing U5M, Ethiopia’s experience provides important lessons in effectively delivering key EBIs in the presence of challenging contextual factors.

## Background

With a population of more than 112 million, Ethiopia is currently the second most populous country in Africa.^1^ About 79% of its population resides in rural areas where communities are typically sparsely distributed.^2^ Health inequities are reflected in notable differences in health outcomes across ethnic-based regions and population groups. Under-5 mortality (U5M), an important area of focus in Ethiopia, decreased from 166 per 1,000 live births in 2000 to 67 in 2016, an impressive decline of about 59%.^3^ However, subnational U5M rates show that this progress was not uniform across the country’s nine regional states and two administrative states. In 2016, U5M rates still ranged widely from 39 per 1,000 live births in Addis Ababa to 125 per 1,000 in the largely pastoralist region of Afar.^3^ There were also differences between residence types, with U5M reported to be about 41% higher in rural areas of the country compared with urban ones.^3^ These inequities mirror differences in coverage of many key health interventions and demonstrate that ensuring equitable access to health services for all is a major challenge in this large and diverse country.^4^

In 1996, the Government of Ethiopia developed a 20-year plan under the Health Sector Development Program (HSDP), which was divided into four series of 5-year plans (HSDP-I – IV). The assessment of HSDP-I revealed that programs were not reaching the grassroots level, and called for mechanisms to improve access. The Ministry of Health (MOH) of Ethiopia invested in expansion and strengthening of its public health system to improve access to primary health care (PHC).^5^ This health system features three tiers – the primary, secondary, and tertiary levels (Figure 1). In rural areas, the primary level includes the PHC unit, each of which consists of five health posts are under one health center, which report to one primary hospital in each *kebele* (Ethiopia’s lowest administrative level). This differs in rural areas, where the primary level simply consists of one health center per *woreda* or district. The secondary and tertiary levels consist of general and six specialized hospitals, respectively.^6^

**Figure 1:**
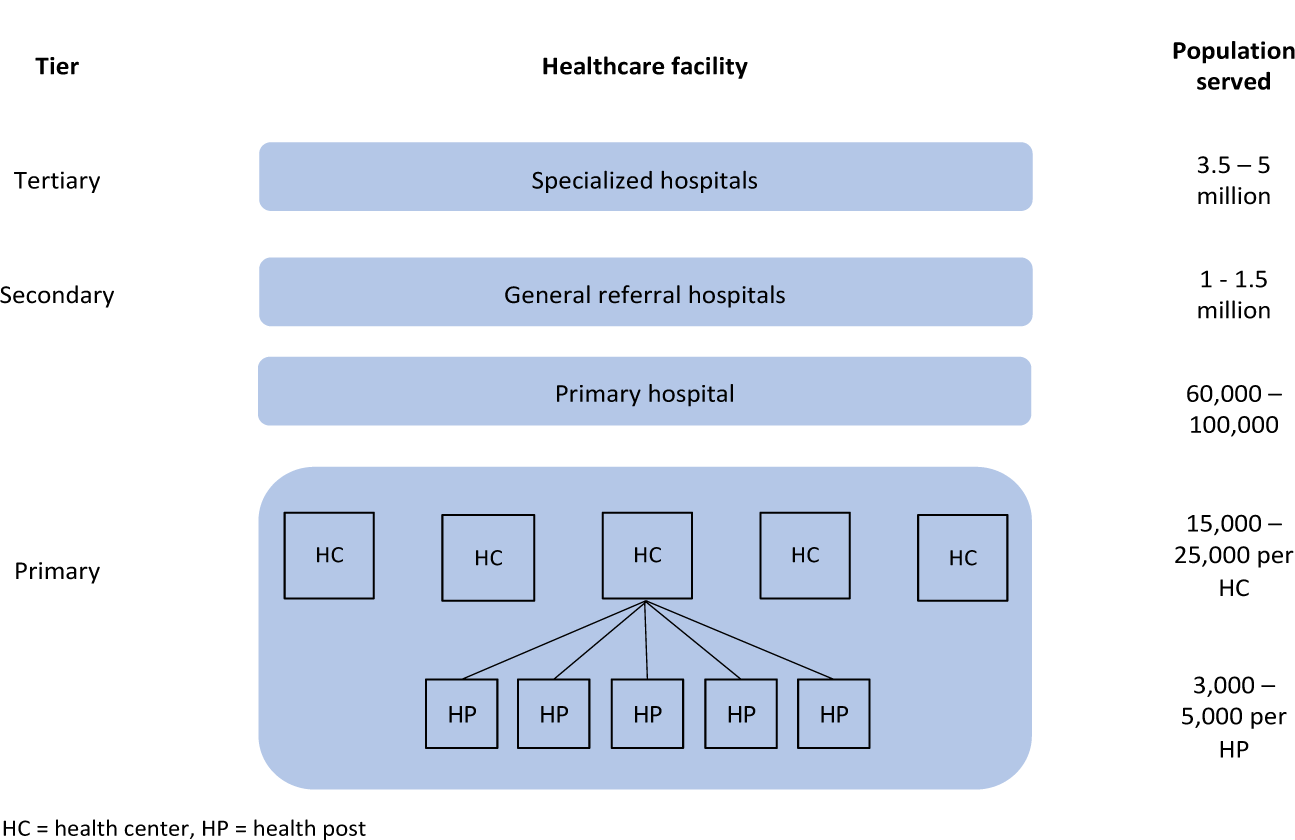
Ethiopia’s three-tiered public health care system

In 2003/2004 the Health Extension Program (HEP) was started to improve coverage and it became part of HSDP-II.^7, 8^ This program was designed to provide community-based health services and address persisting barriers to care in rural areas.^7^ Under the HEP, Ethiopia introduced a cadre of full-time, salaried, and primarily female health extension workers (HEWs), selected by the communities they serve. HEWs complete a one-year formal training after finishing high school, which prepares them to offer an initial package of 16 services at the community level, both at a new facility type called the health post and in homes.^7^ This broad package of services goes beyond the typical scope of community health workers in many countries to include services such as vaccination. In subsequent years following the HEP’s introduction, its scope expanded to improve access to key curative health services, including treatment of diseases like malaria, pneumonia, and diarrhea that represented major causes of death in the country.^7^ By 2009/10, the government had trained and deployed 34,482 HEWs across all regions, with HEW-to-population ratios varying widely by region.^8^ In addition it introduced part-time Health Development Army volunteers in 2011 to support HEWs and assist in health promotion activities.^9^

Implementation research includes the study of how strategies are chosen and used to adopt and integrate evidence-based interventions (EBIs) into real-world settings. It further looks at the contextual factors which serve as facilitators or barriers to improve individual outcomes and population health.^10^ This methodology is of particular importance in improving and sharing lessons from low-and middle-income countries (LMICs) for more effective implementation and improved progress in health outcomes.^11,12^Existing literature has extensively described the role of the HEP in improving coverage of health services through increased access at the community level. For example a systematic review by Assefa et al.^13^ found that the program helped Ethiopia achieve major improvements in several key areas such as communicable diseases, maternal and child health, sanitation and hygiene, community knowledge, and health care seeking.^13^ Other studies have reported on the contribution of the HEP in increasing access to and utilization of health services, particularly for maternal and child health.^14–16^ However, much of the current literature on EBIs is focused on effectiveness and coverage and does not explore what was done, what worked, and implementation strategies utilized.

This paper uses data collected as part of a multi-country implementation research case series to understand successes in addressing amenable U5M deaths through implementation of health system-delivered EBIs between 2000 and 2015. We used case study methodology to understand which implementation strategies Ethiopia utilized alongside leveraging the HEP to implement U5M-targeted health system-delivered EBIs and explore contextual factors that affected the success of the strategies in achieving implementation outcomes. We use the implementation and role of the HEP in two EBIs, the pneumococcal conjugate vaccine (PCV) and integrated community case management (iCCM), to provide a deeper understanding of the processes and outcomes. The results provide transferable lessons for other countries working to accelerate their U5M reduction efforts by leveraging community-based care delivery while targeting inequity in EBI coverage.

## Methods

This study drew from the larger six-country case study series utilizing implementation research methods to understand how countries implemented health-system delivered EBIs known to reduce amenable U5M. Details of the case study methods and our hybrid framework are reported elsewhere.^17^ In brief, this mixed-methods implementation research was informed by a hybrid implementation science framework designed for the project to capture strategies, contextual factors, and implementation outcomes of EBI implementation for reducing U5M in LMICs (Figure 2). This framework included expansion of the Exploration, Preparation, Implementation, Sustainment Framework to include an explicit Adaptation stage (EPIAS).^17, 18^

**Figure 2:**
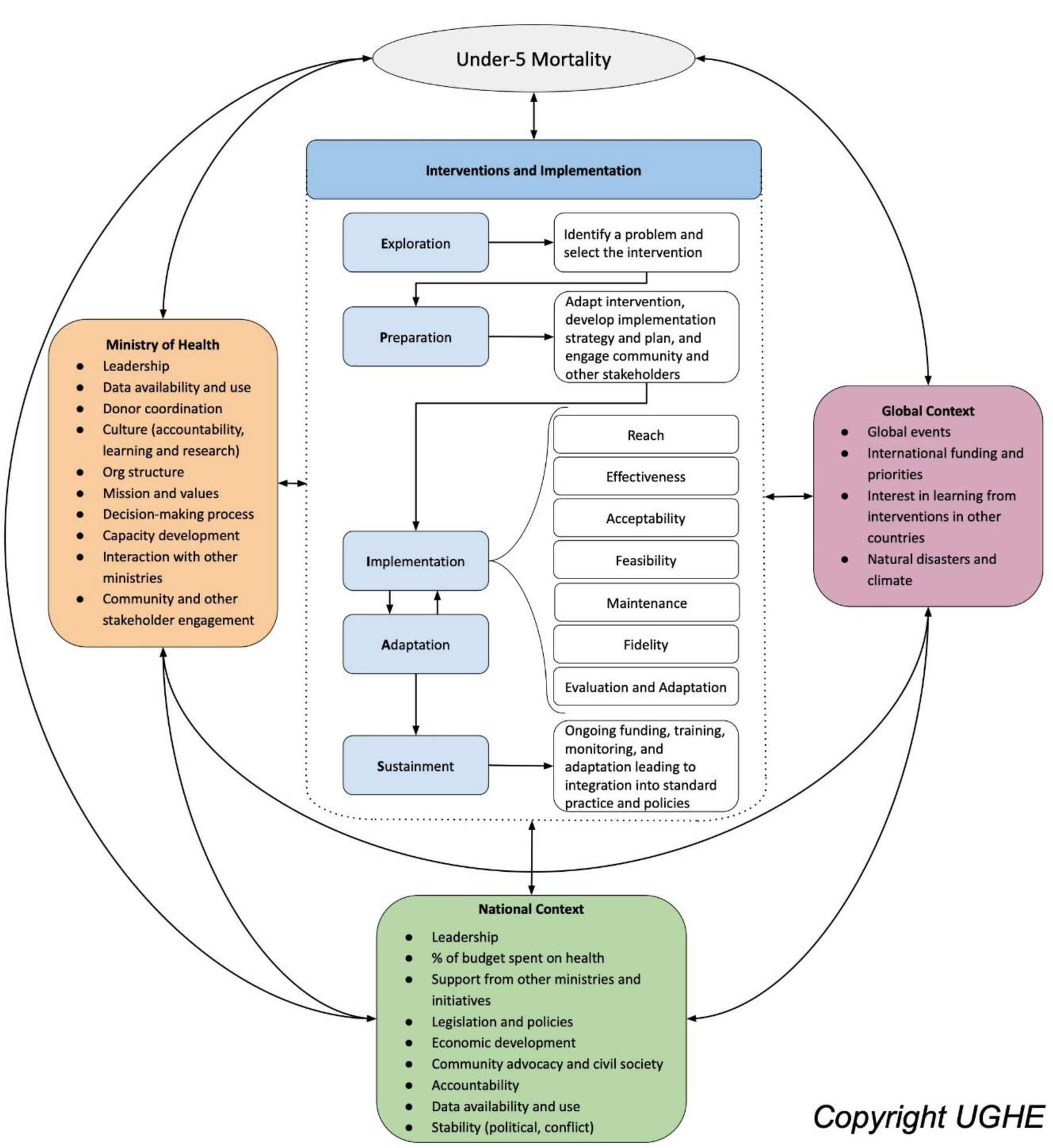
Implementation research framework for understanding evidence-based interventions to reduce under-5 mortality (Hirschhorn, L. R., et al, Gates Open Research, 2021;5,72)

### Data Sources

#### Desk Review

The study team undertook a review of available sources including peer-reviewed and grey literature and program documents focusing on the rates and progress of U5M and implementation of the EBIs known to reduce amenable U5M in countries (supplement Table E1). Initial searches were performed through MEDLINE (PubMed) and Google Scholar using the search terms “child mortality” or “under-5 mortality” and Ethiopia. Further searches included specific EBIs, causes of death, or contextual factors as search terms (e.g. “insecticide-treated nets,” “malaria,” or “community health workers”). Initial desk research was completed by the Strategic Analysis, Research, & Training (START) Center at the University of Washington and the study team expanded on the review through an iterative process throughout the case study as additional data sources or information gaps were identified.

We extracted existing data from sources including the Ethiopian Demographic and Health Survey (DHS, 2001-2016), the World Bank, and WHO/UNICEF to measure changes in EBI coverage, U5M, and trends in health and development indicators related to U5M at the national and subnational levels between 2000 and 2015.

#### Key Informant Interviews

The study team conducted 11 key informant interviews in Ethiopia reflecting a broad range of experience and viewpoints. Key informants (KIs) were chosen based on the targeted EBIs and areas where additional insight was needed from the desk review. We prioritized KIs who were able to provide information on the EPIAS stages during the period of study. Potential KIs were approached by the in-country principal investigator (AT) by telephone or email. Key informants included current and former MOH employees responsible for high-level strategic direction of the ministry or specific disease or intervention areas, implementing partners, and individuals from multilateral or donor organizations who had managed partner-supported activities during the period of interest (2000-2015). We prioritized individuals active in this work during the study period but were also able to capture some experiences from the periods of 1995-1999 and 2017-2019. The interviews were conducted using a semi-structured interview guide based on the hybrid framework. All interviews were conducted in English and led by the project research associates or the in-country principal investigator, who were trained in qualitative interviewing. Interviews were audio recorded and transcribed prior to analysis.

### Data Analysis

We used a mixed-methods Quant-QUAL approach.^18^ Descriptive statistics were used for EBI coverage at the national and subnational levels. We applied the framework to understand the implementation strategies, contextual factors serving as facilitators and barriers at the local, national, and global levels, and implementation outcomes from the literature. *A priori* codes for implementation strategies, outcomes, and contextual factors which were adapted and expanded as emerging codes were identified. The study team used the codes to analyze the transcripts and direct content analysis^19^ was used to identify strategies, contextual factors, and implementation outcomes. Qualitative and quantitative data were triangulated to link the strategies, contextual factors, implementation outcomes, and coverage, and to emerge transferable lessons for other countries learning from EBI implementation in Ethiopia.

## Results

### Role of Health Extension Program

We found from the desk review and key informant interviews that for the majority of the targeted EBIs implemented in Ethiopia to reduce U5M the HEP was utilized as an important implementation strategy (Table 1). Ethiopia frequently leveraged the HEP as a platform for delivery of new services, with HEWs playing an important role as key implementers. Service delivery by HEWs increased coverage of many EBIs compared to implementation at health facilities alone. HEWs made health services more accessible to rural and pastoralist communities that previously had low access to health facilities and services, a contextual factor that served as a barrier to high coverage of health services and interventions. This strategy of leveraging the HEP improved feasibility and reach of introduction and enabled scale up of interventions in more resource-limited and rural settings in Ethiopia by using the strong, widespread platform. Utilization of the HEP also helped Ethiopia ensure acceptability of new interventions since service delivery involved HEWs already working in communities. The strategy of embedding new interventions into the HEP additionally improved the sustainability of new interventions by building upon an existing service delivery platform rather than relying on new, vertical ones.

**Table 1:** *List of Evidence-Based Interventions Studied and Use of Health Extension Program*

| Area of focus | Evidence-based intervention | Health Extension Program utilized |
| --- | --- | --- |
| Pneumonia<br>Diarrhea<br>Malaria | Facility-based integrated management of childhood illness (FB-IMCI) | x |
|  | Integrated community case management (iCCM) | x |
|  | Oral rehydration solution (ORS) and zinc | x |
|  | Rotavirus vaccination | x |
|  | Pneumococcal vaccination | x |
|  | *Haemophilus influenzae* type B vaccination | x |
|  | Insecticide-treated nets | x |
|  | Indoor residual spraying | x |
| Other vaccine-preventable diseases | Measles vaccination | x |
|  | Meningococcal vaccination | x |
| Human immunodeficiency virus (HIV) | Prevention of mother-to-child transmission of HIV | x |
|  | Early infant diagnosis and treatment of HIV | x |
| Nutrition | Management of severe acute malnutrition | x |
|  | Vitamin A supplementation | x |
| Neonatal period | Improving access to antenatal care | x |
|  | Management of pregnancy-associated hypertensive disorders |  |
|  | Maternal tetanus vaccination |  |
|  | Facility-based delivery and skilled birth attendance | x |
|  | Basic and comprehensive obstetric and newborn care |  |
|  | Caesarean section |  |
|  | Clean delivery practice | x |
|  | Corticosteroids for preterm labor |  |
|  | Active management of delivery |  |
|  | Neonatal resuscitation |  |
|  | Neonatal intensive care units |  |
|  | Kangaroo mother care |  |
|  | Management of neonatal sepsis | x |
|  | Post-partum visits | x |

In addition to direct service provision, HEP activities were designed to increase demand and uptake of maternal and child health services based at health facilities. Strategies included regular household visits, conducting community engagement and health promotion activities including sensitization to promote facility-based delivery, an EBI with very low coverage historically. Nationally, the Women Development Army cadre of volunteers were introduced into the HEP in 2011 to enhance the HEP’s ability to reach households. They also helped strengthen HEP activities and improve utilization of services through demand generation activities and linking community members and HEWs.^9^

We identified several other implementation strategies to introduce and scale up EBIs to reduce U5M. The most common implementation strategies Ethiopia utilized (shown in Table 2 as used for PCV and iCCM) included: national policy and development planning, leveraging and coordinating strong donor and partner support, using data for evidence-based decision-making, and integration into or leveraging of existing programs or platforms.

**Table 2:** Selected Contextual Factors, Implementation Strategies, and Role of Health Extension Program

| **Evidence-based intervention** | **Contextual factors** | **Implementation strategies utilized** | **Role of Health Extension Program (HEP)** |
| --- | --- | --- | --- |
| Pneumococcal conjugate vaccine | • Donor resources (+)<br>• Investment in health (+)<br>• Strong community health system (+)<br>• Ongoing health systems strengthening efforts (+)<br>• Data availability and use (+)<br>• Geography and inequity of coverage across regions (-)<br>• Limited vaccine and resource availability (-) | • Data use for decision-making and local data generation<br>• Government co-financing<br>• Stakeholder engagement<br>• Community engagement and social mobilization activities<br>• Introduction via phased approach prioritizing high-risk areas<br>• Utilization of donor and partner resources<br>• Systems assessment and strengthening<br>• Adaptation for local context<br>• Training and supportive supervision (cascaded)<br>• Mass vaccination campaigns with follow-up campaigns every five years<br>• Integration into routine vaccine schedule<br>• Translation of advocacy, communication, and educational materials into local languages | • Vaccine provided by HEWs at health posts as part of routine vaccination<br>• Implementation of community engagement and social mobilization activities in communities |
| Integrated community case management (iCCM) | • Effective national leadership and control (+)<br>• Donor and implementing partner resources (+)<br>• Strong community health system (+)<br>• Data availability and use (+)<br>• Geography and inequity of coverage across regions (-)<br>• Low utilization of health services (-)<br>• Supply chain (-) | • Integration into existing program<br>• Leveraging and coordination of donor and partner support<br>• Data use for decision-making<br>• National policy and development planning<br>• Utilization of supportive supervision<br>• Adaptive management<br>• Monitoring and evaluation<br>• Decentralization and standardization of training curriculum within existing HEW training program<br>• Quality improvement activities<br>• Adaptation to include major causes of death and meet needs of neonatal mortality<br>• Initial introduction in four regions | • iCCM implemented within existing HEP<br>• Care delivery by HEWs at health posts<br>• iCCM integrated into pre-service training for HEWs<br>• Use of health development army volunteers to generate demand for iCCM services and link community members with HEWs for care |

(+) = facilitator, (-) = barrier; HEW = health extension worker

The HEP’s role in the implementation of two EBIs, PCV and iCCM, is further described below. These two EBIs were introduced and successfully scaled to a national level during the study period to address major causes of U5M and are discussed to illustrate the role of the HEP in implementation and delivery of vaccines and curative care in Ethiopia.

### Pneumococcal Conjugate Vaccine

The PCV-10 vaccine was introduced in late 2011 to prevent severe forms of pneumococcal disease such as pneumonia and meningitis that were previously estimated to account for up to 28% of all deaths among children under 5 in Ethiopia.^20^ The work followed the implementation pathway of the EPIAS framework (Table 2). Exploration was followed by an intensive preparation stage typical of new vaccines in the country that included implementation strategies such as obtaining procurement support from Gavi, the Vaccine Alliance. This was leveraged through a new Pneumococcal Advance Market Commitment initiative (which served as a facilitating contextual factor by improving availability of funds targeting the new vaccine), comprehensive cascade training of health workers, and adoption of training manuals. During the preparation phase, HEWs also aided in implementation of community mobilization and advocacy activities, important implementation strategies to create demand for these vaccines.^21^ The strategy of leveraging the HEP as a platform for service delivery allowed Ethiopia to successfully implement and scale PCV-10, supported by these other strategies. In addition to mass vaccination campaigns, a catch-up strategy to vaccinate all children under the age of 1 year was used, and the vaccine was provided through routine, free vaccination services at health facilities, including by HEWs at health posts. This provision of routine immunization services at health posts allowed new vaccines such as PCV-10 and others (such as rotavirus and pentavalent vaccines) to quickly achieve widespread implementation at a national scale and reach rural communities.

PCV was introduced throughout the country simultaneously and with high levels of general acceptability of vaccines in the community, according to interviewees in a 2015 study.^22^ By 2014/15, WHO/UNICEF estimated coverage of all three doses of PCV-10 to be 76%.^23^ Despite rollout in all regions and implementation strategies targeting national reach, geographic coverage of the vaccine was not equitable, with large regional differences.^3^ According to a key informant, these inequities reflected barrier contextual factors in differences in health system strength and governance between the regions, as well as presence of pastoralist populations in regions such as Afar and Somali. These two largely pastoralist regions had the lowest coverage of three doses of PCV reported by the 2016 DHS – 23.6% and 22.9%, respectively.^3^ We found that other contextual factors such as limited vaccine and resource availability were also hindering factors. Conversely, availability of donor resources, government investment in health, ongoing health systems strengthening efforts, and data availability and use were facilitating factors.

### Integrated Community Case Management of Newborn and Childhood Illness

Another important EBI introduced during the study period was community-delivered integrated management of childhood illness (IMCI), or integrated community case management (iCCM). Ethiopia previously introduced the facility-based IMCI program to address several leading causes of death in children under 5 in an integrated manner at health centers. However, the government found that access to curative child health services remained lower than expected after scale-up. In response, the MOH began adding selected curative services to the scope of existing HEWs for a more community-based delivery strategy. Ethiopia’s national iCCM program formally launched in early 2010 within the HEP following pilot studies conducted by partners, an implementation strategy that generated local evidence on feasibility, and extensive preparation in collaboration with several partners. As part of iCCM, HEWs provide community-level management of malaria, diarrhea, pneumonia, malnutrition, and, later, essential newborn care and management of common neonatal problems including neonatal sepsis at health posts.

Similar to many other EBIs implemented in Ethiopia, the program utilized a strategy of phased introduction after the pilot testing to improve feasibility of expansion given Ethiopia’s geography, which was an important barrier contextual factor. ICCM was first implemented at a small scale in two of Ethiopia’s nine regions and eventually scaled to the entire country. Four agrarian regions were initially targeted (Amhara, Oromia, Southern Nations, Nationalities, and People’s Region (SNNPR), and Tigray) due to their greater population density and strength of HEP implementation compared with other regions. It was later scaled up throughout the country, including in pastoralist regions after development of a contextualized implementation guide for pastoralist areas. By 2014, iCCM was implemented by almost 30,000 HEWs working at 14,500 health posts in eight regions, representing 86% of the country’s districts.^24^ Notably, the program was not rolled out in the country’s urban areas or two city administrations, Addis Ababa and Dire Dawa.

Though implementation of iCCM had a significant impact on geographic access to curative child health services in rural areas of the country, we identified important challenges to the program that likely limited coverage, and ultimately the program’s benefits. One significant barrier contextual factor affecting coverage of services was health-seeking behavior in many areas of the country. Home visits by HEWs as part of iCCM enabled early identification of sick children and improved care-seeking behaviors; care-seeking for children with ARI symptoms and diarrhea increased by 48% and 112% respectively from 2000 to 2016. ICCM introduced evidence-based management of common childhood illnesses at the community level. However, uptake remained limited, with fewer than 50% of children under 5 with acute respiratory infection symptoms or diarrhea taken to any health facility, including a health post manned by HEWs, in 2016.^3^ The 2014 Ethiopia Service Provision Assessment Plus cross-sectional survey identified additional capacity issues at health posts affecting provision of iCCM.^25^ While availability of curative services for children under 5 was very high at health posts, overall quality of care and adherence to iCCM guidelines was found to be poor. Treatment may have been affected by weak supply chain with low availability of essential medicines required for iCCM services – zinc for treatment of diarrhea and antibiotics were both available in fewer than half of health posts assessed.^26^ In response to stock outs of essential medicines, the program later adapted its supply chain management to use a “pull” supply chain system, or integration into the Integrated Pharmaceutical Logistic System (IPLS), that improved access to drugs at the health post level. According to a key informant, “*this adaptation minimized drug wastage, improved access to drugs at the health post level, and improved equity.”*

Despite these challenges, we found evidence of improved care-seeking and coverage and equity of treatment of the conditions by the iCCM program addressed following implementation. While overall coverage increased across groups, some inequities remained. Though these services were offered for free at the health post and health center level, mitigating some financial barriers to care, children in the highest wealth quintile with diarrhea were more likely to be taken to a health facility (61%) than those in the lowest and second quintiles (40%) (Table 3b). Oral rehydration solution (ORS) for treatment of diarrhea, a service included in iCCM, doubled over the same period. A notable increase in coverage of ORS in rural areas (Table 3b) reflected overall improvement in equity. ORS use increased in all regions except Somali, where coverage declined. Despite improvement in most regions, regional differences in coverage persisted, ranging from 23.9% in Oromia to 52.5% in Benishangul-Gumuz in 2016 (Table 3b).^3^

**Table 3a:** Coverage of Integrated Community Case Management (iCCM) Interventions by Region

| Intervention | Addis Ababa |  | Afar |  | Amhara |  | Ben.-Gumuz |  | Dire Dawa |  | Gambella |  | Harari |  | Oromia |  | Somali |  | SNNPR |  | Tigray |  | National |  |
| --- | --- | --- | --- | --- | --- | --- | --- | --- | --- | --- | --- | --- | --- | --- | --- | --- | --- | --- | --- | --- | --- | --- | --- | --- |
|  | 2000 | 2016 | 2000 | 2016 | 2000 | 2016 | 2000 | 2016 | 2000 | 2016 | 2000 | 2016 | 2000 | 2016 | 2000 | 2016 | 2000 | 2016 | 2000 | 2016 | 2000 | 2016 | 2000 | 2016 |
| U5 with ARI symptoms taken to facility (%) | 58.4 | . | 21.9 | 53.1 | 11.5 | 28.1 | 28.0 | . | 33.1 | . | 41.3 | . | 40.5 | . | 16.6 | 30.2 | 49.4 | . | 15.4 | 45.4 | 12.3 | 36.7 | 21.3 | 31.6 |
| U5 with diarrhea taken to facility (%) | 31.1 | 54.4 | 15.4 | 58.9 | 10.8 | 41.0 | 25.2 | 64.7 | 25.9 | 66.4 | 31.4 | 65.4 | 29.9 | 61.5 | 12.5 | 45.9 | 42.3 | 45.5 | 13.2 | 50.5 | 17.1 | 52.6 | 21.1 | 44.8 |
| ORS for diarrhea treatment (%) | 40.1 | 49.1 | 23.6 | 35.8 | 9.0 | 24.1 | 13.4 | 52.5 | 25.9 | 50.6 | 32.6 | 43.3 | 26.2 | 44.3 | 12.9 | 23.9 | 50.5 | 46.2 | 13.0 | 34.3 | 15.3 | 43.5 | 13.1 | 19.5 |

ARI = acute respiratory infection, ORS = oral rehydration solution, U5 = under-5

**Table 3b:** Coverage of Integrated Community Case Management (iCCM) Interventions by Sex, Residence, and Wealth Quintile

| Intervention | Sex |  |  |  |  |  | Residence |  |  |  |  |  | Wealth Quintile |  |  |  |  |  |
| --- | --- | --- | --- | --- | --- | --- | --- | --- | --- | --- | --- | --- | --- | --- | --- | --- | --- | --- |
|  | Male |  |  | Female |  |  | Urban |  |  | Rural |  |  | Lowest |  |  | Highest |  |  |
|  | 2000 | 2016 | % change | 2000 | 2016 | % change | 2000 | 2016 | % change | 2000 | 2016 | % change | 2000 | 2016 | % change | 2000 | 2016 | % change |
| U5 with ARI symptoms taken to facility or provider (%) | 22.3 | 34.1 | +54.3 | 20.2 | 29.1 | +44.1 | 48.1 | 64.4 | +33.9 | 19.3 | 29.2 | +51.3 | 11.5 | 25.0 | +117.4 | 39.3 | 44.2 | +12.5 |
| U5 with diarrhea taken to facility (%) | 23.6 | 41.5 | +75.8 | 18.4 | 48.4 | +163.0 | 48.4 | 60.9 | +25.8 | 18.9 | 42.9 | +127.0 | 22.1 | 40.1 | +81.4 | 39.2 | 61.1 | +55.9 |
| ORS for diarrhea treatment (%) | 13.3 | 30.2 | +127.1 | 12.8 | 28.8 | +125.0 | 47.3 | 40.5 | -14.4 | 10.3 | 28.3 | +174.8 | 8.2 | 26.7 | +225.6 | 33.2 | 41.4 | +24.7 |

*ARI = acute respiratory infection, ORS = oral rehydration solution, U5 = under-5*

### Implementation Strategies and Contextual Factors

Ethiopia’s successful introduction and national scale-up of key EBIs to reduce U5M utilized various implementation strategies. One commonly used strategy was leveraging existing programs such as the HEP to implement new EBIs, an approach that improved feasibility, reach, and sustainability of new EBIs given the country’s limited resources and geography. Alongside this approach, Ethiopia used several other implementation strategies, including using data for evidence-based decision-making, leveraging donor and partner resources, introducing EBIs via a phased approach, and national policy and development planning. Implementation was also impacted by cross-cutting contextual factors that served as facilitators and barriers. In Ethiopia, we found the presence of a strong community health system, other health systems strengthening efforts, data availability and use, and availability of donor and implementing partner resources were contextual factors that facilitated U5M reduction efforts. We found that, meanwhile, other contextual factors such as geography, supply chain issues, and low utilization of health services served as barriers. For many EBIs, pastoralist culture was a hindering factor affecting coverage in regions including Afar, where the majority of the population is pastoralist and U5M rates are some of the highest in the country.

## Discussion

We found that most of the EBIs implemented in Ethiopia to reduce U5M between 2000 and 2015 leveraged the HEP as an implementation strategy, using the program as a platform for delivery of new services. Ethiopia also used a number of other strategies to successfully implement and scale up many of the existing and new EBIs, although gaps remained in coverage and in fidelity.

Prior studies have reported on the contribution of the HEP in increasing access and uptake of health services, including maternal and child health.^13–16^ They have also found that the program helped Ethiopia achieve improvements in areas such as maternal and child health, communicable diseases, and sanitation and hygiene.^13^ The contributions of this paper allow us to better understand some of the mechanisms through which this strategy worked and other implementation strategies that were used alongside the HEP to implement EBIs and contextual factors that affected their success. For example, implementation strategies such as use of data for evidence-based decision-making and leveraging donor and partner resources were beneficial in introducing and scaling up interventions. Implementation of PCV and iCCM were impacted, however, by contextual factors such as geography, supply chain challenges, and care-seeking behavior.

Similar to our findings on iCCM, a study on the assessment of success and challenges of HEP in Ethiopia between 2003 to 2018 found readiness and availability of services including staffing, equipment, and adequate supplies at the health post level where the HEP operates and service utilization were key barriers for the successful implementation various EBIs by HEWs.^13^

Literature on CHW programs in other LMICs is abundant. As in Ethiopia, CHW programs in countries including Brazil, Nepal, and Iran have increased access to health services in rural areas and contributed to improved outcomes in these areas.^27–30^ CHW programs often share the HEP’s challenges, such as those related to supply chain in Zambia and Pakistan or community care-seeking preferences in India.^30–,32^ Literature shows comparative strengths of the HEP that have likely contributed to the program’s success. For example, Accredited Social Health Activists (ASHAs) in India conduct activities like that of HEWs but are considered part-time volunteers. While HEWs are full time and salaried, ASHAs are reported to be dissatisfied with remuneration as they are only paid for a select few interventions in their wide scope.^32^ CHW training in some countries such as Pakistan and Zimbabwe has been reported to be inadequate, though this is a strength in Ethiopia where HEWs receive more than a year of pre-service training.^31, 33^

Ethiopia provides a valuable example for other low-resource settings in its utilization of a community health program to improve access to both preventive and curative EBIs. Some countries have not yet adopted a similar approach of utilizing CHWs to provide very decentralized and comprehensive services at the community level. Ethiopia’s implementation strategies demonstrate the importance of building upon existing, widespread programs to introduce new EBIs more feasibly at a large scale or to further expand access and reach of existing ones. It has been well documented that the HEP enabled Ethiopia to improve critical health outcomes in areas of maternal and child health, communicable diseases, hygiene and sanitation, knowledge, and increasing health care-seeking behavior.^13^

The experience of Ethiopia also demonstrates the importance of efforts to adapt strategies or introduce new ones to encourage uptake and ensure equity of coverage of services offered at the community level reflecting subnational variability in geography, culture, and other contextual factors. Establishment and expansion of the HEP was a key strategy to improve access to health services at the community level, particularly in Ethiopia’s vast rural and pastoralist areas with historically low access to health facilities. There is also evidence of pro-poor public spending on health in Ethiopia which supported the implementation of EBIs by the HEP.^13^ The Ethiopian government allocated nearly 60% of health expenditures at health centers which supports the work of the HEP.^34^

The challenges we identified in implementation were similar to contextual barriers described elsewhere. A systematic review of the HEP similarly highlighted the role of the HEP in improving maternal and child health services at the health center and community levels.^13^ However, remaining challenges include capacity of health posts related to supplies, variation in performance of HEWs across geography, work overload, and contextual factors related to the wider health system issues.^13^ For facility-based services, HEWs played a role in promoting care-seeking. A similar role was adopted by Health Development Army volunteers in 2011 as an adaptation to the HEP. Since uptake may be related to perceived quality of community-level care, ensuring high quality of care within community health programs should be a priority for countries leveraging them. Ethiopia used a system of supportive supervision during implementation of many EBIs to improve fidelity, a strategy that may be valuable in many settings. However, quality of care remains an area for improvement that became a priority area for the national Health Sector Transformation Plan.^35^ Coverage of many EBIs, even ones such as ORS with expanded community-level access during the study period, still varies greatly by region, residence type, and wealth quintile in Ethiopia. This is an ongoing challenge that other countries are likely to face and should consider during planning and preparation of implementation.

Our study must be interpreted in light of its limitations. This case study is limited by the fact that publicly available data, reports, and publications often do not describe the implementation outcomes of interest at a subnational level, and data often are either reported at a national level or for a specific subnational population. This was a retrospective study that relied on recall for critical qualitative components, with the corresponding risk of recall bias.

### Conclusions

As LMICs work towards further reducing child mortality, existing community health programs represent a valuable opportunity for introduction and expansion of new EBIs. Ethiopia utilized a strong, widespread community health program, the Health Extension Program, to implement new EBIs, including the pneumococcal vaccine and integrated community case management. Leveraging the HEP as a platform for service delivery allowed Ethiopia to successfully introduce and scale new and existing EBIs nationally. Further effort is required to reduce the equity gap in coverage of EBIs among pastoralist and rural communities. Ethiopia’s work in utilizing the HEP alongside other implementation strategies to effectively deliver key EBIs in the presence of potentially challenging contextual factors provides important lessons for other countries aiming to reduce under-5 mortality.

### Declarations

### Ethics approval and consent to participate

The study was reviewed by the Institutional Review Board in Ethiopia (Approval number PM23/281). All KIs provided informed consent before interviews were conducted. The overall project was also reviewed by the Rwanda National Ethics Committee and Northwestern University and determined to be non-human-subjects research.

## Consent for publication

Not applicable

### Data availability

The datasets used and/or analyzed during the current study are available from the corresponding author on reasonable request.

## Competing interests

The authors declare that they have no competing interests.

## Funding

This work was completed as part of a larger case study series funded by the Bill & Melinda Gates Foundation and Gates Ventures. These funding bodies were not directly involved in the development of this manuscript.

### Authors’ contributions

LD, AMT, AB, and LRH made substantial contributions to the design of this work. LD, AA, JTN, AMT, AB, and LRH contributed to the data analysis. LD, AA, AMT, MZ, AT, YT, SG, YS, JTN, AV, AB, and LRH interpreted the results to form the manuscript. LD, AA, AMT, MZ, AT, YT, SG, YS, JTN, AV, AB, and LRH contributed to writing and revision of the manuscript. All authors have approved the final version.

## Data Availability

https://data.worldbank.org/indicator/SP.URB.TOTL.IN.ZS?locations=ET

https://www.statcompiler.com/en/

## Acknowledgements

We would like to acknowledge and thank the key informants and other stakeholders in Ethiopia who provided essential information, historical perspectives and narratives, and feedback on our findings, ensuring we captured accurate reflection possible on Ethiopia’s journey to reducing under-5 mortality.

## Notes

### Competing Interest Statement

The authors have declared no competing interest.

